# Predicting Clinical Course from Subcortical Shape in Provisional Tic Disorder

**DOI:** 10.1101/2021.11.04.21265815

**Authors:** Tiffanie Che, Soyoung Kim, Deanna J. Greene, Ashley Heywood, Jimin Ding, Tamara Hershey, Bradley L. Schlaggar, Kevin J. Black, Lei Wang

## Abstract

The NewTics study examined children at the onset of tic disorder (with tics for less than 9 months: NT group), a population on which little research exists. Here, we investigate relationships between the shape of subcortical nuclei and tic symptom outcomes. 187 children were assessed at baseline and a 12-month follow-up: 88 with NT, 60 tic-free healthy controls (HC), and 39 with chronic tic disorder or Tourette syndrome (TS), using T1-weighted MRI and total tic scores (TTS) from the Yale Global Tic Severity Scale to evaluate symptom change. Subcortical surface maps were generated using FreeSurfer-initialized large deformation diffeomorphic metric mapping, and linear regression models were constructed to correlate baseline structural shapes with follow-up TTS while accounting for covariates, with relationships mapped onto structure surfaces. We found that the NT group had a larger right hippocampus compared to healthy controls. Surface maps illustrate distinct patterns of inward deformation (localized lower volume) in the putamen and outward deformation (localized volume gain) in the thalamus for the NT group compared to healthy controls. We also found distinct patterns of outward deformation in almost all studied structures when comparing the TS group to healthy controls. In the significant vertices of this comparison, the caudate further exhibited an overall trend of greater outward deformation (compared to the template) in the TS group compared to both the NT group and controls. When comparing the NT and TS groups, the NT group showed consistent outward deformation in the caudate, accumbens, putamen, and thalamus. Since the NT group has had tics only for a few months, we can rule out the possibility that these subcortical volume differences are caused by living with tics for years; they are more likely related to the cause of tics. Subsequent analyses including clinical symptoms revealed that a larger pallidum and thalamus at baseline predicted less improvement of tic symptoms in the future. These observations constitute some of the first prognostic biomarkers for tic disorders and suggest that these subregional volume differences may be associated with outcome of tic disorders.

## Introduction

Persistent (chronic) tic disorders (CTD) were once thought to be rare but are now known to affect a substantial number of elementary school children [1]. Tics are sudden, repetitive, nonrhythmic movements or vocalizations such as blinks or grunting [2]. Transient tics affect at least 20% of children, though only about 3% of all children have tics for a full year, the requirement to diagnose a chronic tic disorder or Tourette syndrome (TS/CTD) [3, 4]. When tics are present but less than a year has passed since the first lifetime tic, Provisional Tic Disorder is diagnosed [1]. Efforts to identify biomarkers for tics and study the pathophysiology behind tic disorders have recently been increasing, although our understanding is still limited [5, 6].

A paucity of available autopsy data elevates the importance of *in vivo* neuroimaging in studying the pathophysiology of tic disorders. Studies exploring differences in subcortical structure and function have often been contradictory, with some finding no significant differences in basal ganglia volumes or shape between children with TS and matched control children [6]. Two groups found greater putamen volume in TS compared to HC, but a larger study found smaller volume [7-9]. A large study of basal ganglia volume found the caudate to be 4.9% smaller in the TS group compared to tic-free children [10]. Smaller studies also found lower caudate volume [11, 12], but another large pediatric study identified no difference between TS and control groups [13].

Previous such studies have also focused only on TS (diagnosed chronic tics) and control samples. Therefore, we cannot determine whether the identified differences reflect an underlying cause of tics or secondary changes due to prolonged tic presence. Examining children at the onset of tic symptoms will more likely lead to identifying biomarkers related to the primary cause of tics.

The ongoing NewTics study examined children who had tics for less than 9 months (new tics, or NT) [14, 15]. Previously, little research existed on this population, there were even fewer results on prognosis, and those had been contradictory [4]. The NewTics study tested whether features including subcortical structures measured shortly after tic onset could predict symptom severity 12 months after tic onset. A previous volumetric MRI analysis using data from 65 children with NT (a subset of the current sample) found that striatal volumes did not predict outcome, but a larger hippocampus at baseline predicted worse severity at follow-up [16]. However, using whole-structure volume estimates alone may yield false negative findings by overlooking local deformities in shape.

In the present study, we further investigated neurobiological characteristics and predictors of tic disorders by examining relationships of the *shape* of these subcortical structures with tic symptom outcomes (NT) and with diagnosis (NT, TS, and tic-free controls). Three-dimensional surface analysis can detect subtler or more localized volumetric changes that are not revealed in whole-structure, scalar volumetric analysis [17]. Previously, diffeomorphic mapping of structural magnetic resonance imaging (MRI) has successfully mapped pathological biomarker patterns onto surface-based representations of anatomical structures [18]. We also take advantage of a much larger data set, a superset of the sample previously studied with scalar volumetrics [16].

We predicted that baseline volumes would differ across NT, TS, and control groups, and that subcortical shape would demonstrate distinct patterns of shape deformation in tic disorders. We further predicted that we would find distinct regions of shape deformation in subcortical structures at baseline that predict clinical outcome in terms of tic severity changes 3-12 months later. We also tested whether shape deformation analyses would confirm the previous finding that hippocampal volume predicted symptom severity outcome using a 3D method in an expanded sample.

## Materials and Methods

### Subjects and data collection

Subjects. The sample consisted of 187 children across 3 groups: children examined within 9 months after tic onset (median 3.5 months; new tic, or NT), tic-free children with no parental or sibling history of tics (healthy controls, HC), and children who at the time of screening already have TS/CTD (TS group) [14]. NT children returned for clinical evaluation at the one-year anniversary of the best estimate of tic onset. For TS and HC children, follow-up visits occurred as near as possible to the same time after screening as it did for their matched NT child, based on age, sex, and handedness. Healthy controls were collected across different studies/sources. Detailed information about recruitment methods appeared elsewhere [14].

Enrollment criteria. Participants were ages 5-15 at enrollment (ages 5-10 after 3 children older than 10 were enrolled). NT children had current tic symptoms, with the first tic starting less than 9 months before enrollment. Exclusion criteria are detailed elsewhere [14]. The TS group included children who met DSM-5 criteria for TS/CTD at enrollment. The exclusion criteria used reflect those of the NT group.

Control children were confirmed to have no tics after thorough history from child and parent separately, neurological examination, and at least 10 minutes of observation via video while the child sat alone.

Clinical data collection. For participants exhibiting tic symptoms, a best-estimate date of onset was recorded after careful inquiry as described in [19]. Total tic scores from the Yale Global Tic Severity Scale (YGTSS), reflecting current tic severity, were determined during a neurological and psychiatric examination performed by author KJB, supplemented with remote video observation of the participant sitting alone. Additional assessments including a K-SADS diagnostic interview and measures of OCD and ADHD symptoms were done at the time of screening [14]. The Total Tic Score (TTS) comprises half of the YGTSS score and has a range of 0-50; a higher score indicates more severe tic symptoms [20].

Imaging data collection. All subjects at entry were scanned with 1 mm^3^ T1-weighted magnetization prepared rapid gradient echo (MPRAGE) and T2-weighted sequences using three different 3T scanners across the data acquisition period. Details of scan parameters are given in Kim et al (2020) [16]. About half of the scans (the newer ones) were acquired with a prospective motion correction sequence (vNavs) [21].

Additional measures were taken prior to scanning to reduce motion effects on images, including training in a mock scanner, an informational video, and a game for children to practice holding still during scanning, adapted from a previous study in children [22].

### Data processing / Image processing

Data processing began with the use of the FreeSurfer (version 6.0.0) software’s probabilistic voxel-based classification, which provided initial subcortical segmentations [23]. Surfaces of the hippocampus, amygdala, basal ganglia (caudate, putamen, pallidum, and nucleus accumbens), and thalamus were automatically generated for each participant using multi-atlas FreeSurfer-initialized large deformation diffeomorphic metric mapping (FS+LDDMM), which utilizes automated brain segmentations based on multiple template images and allows for image alignment and intensity normalization to produce smooth transformations for each region of interest [23, 24]. Combining maps from multiple atlases that best match an individual’s scan features has shown improved segmentation accuracy and reduced biases [25]. An experienced rater (the first author) inspected the final surfaces and made minor manual edits to the initial segmentation on poor surface maps of 6 subjects. The edited segmentations were then re-processed via LDDMM to yield accurate maps to be included in subsequent surface analyses.

To equalize the effect of total brain volume across participants, surfaces were then scaled with a scale factor calculated for each subject using the population total intracranial volume (TIV), the individual’s TIV, and the voxel size of their scan. To calculate the (one-dimensional) scale factor, we used the formula: (Population TIV / Individual TIV)^⅓^ × Voxel Size.

Local shape variation for each participant was calculated from the population average of all participants by quantifying the vertex-to-vertex perpendicular change between surfaces, which were assigned a positive (outward variation from the population average) or negative (inward variation from the population average) value [23]. Subcortical volumes for each participant were determined using the volume enclosed within the surfaces. FreeSurfer also reported estimated total intracranial volume (TIV) to be used as a covariate in surface analyses, as TIV estimated by FreeSurfer segmentation is correlated with subcortical volumes [24].

### Statistical analysis

#### TTS

Paired T-tests were conducted to determine the significance of TTS changes from baseline to 12 months within each group, as well as between the NT and TS groups.

#### TIV

We first conducted a one-way ANOVA on TIV (dependent variable) to determine group (independent variable) effects with post hoc Tukey HSD tests for pairwise differences, with and without age and sex as covariates. The tests showed significant group effects (see results); therefore, TIV was used as an additional covariate in all subsequent analyses.

#### Scanner type

We first conducted a chi-square test that determined that scanner types did not differ between groups. Additionally, we conducted one-way ANOVA tests to determine the effects of scanner type on structural volumes. We found that the subcortical structures did not differ in volume based on the scanner type, and subsequent ANCOVA and post hoc tests found that group differences found in TIV between NT and healthy controls still occur after controlling for the scanner type. Thus, we did not use scanner as a covariate in subsequent analyses.

#### Race

The same tests done for scanner type (see above) were applied to race as well, and similar results were found. We thus also did not use race as a covariate in subsequent analyses.

#### Subcortical volumes

Using the surface-defined volumes for each subcortical structure, we first found estimated marginal means and standard errors for each group. ANCOVA and post hoc tests on subcortical volumes (dependent variables) were then conducted to compare group (independent variable) differences of baseline structural volumes, using age and sex as covariates. All structures were examined by combining left and right subcortical structures, since we did not have a lateralized hypothesis. If groups were found to differ significantly for a specific structure, further analyses were conducted to examine left and right structures separately. Analyses run in R used version 4.0.5, with psych and ggplot2 packages [26, 27].

We then performed a partial correlation analysis using baseline structural volume to predict 12-month TTS in the NT group while controlling for TTS at screening, age, and sex.

#### Shape

For group comparisons, ANCOVAs were conducted to compare pairwise group differences of baseline surface shape (i.e., NT vs. HC, TS vs. HC, and TS vs. NT). All models included covariates for age and sex. Surface comparisons were conducted using SurfStat implemented in MATLAB [28]. This software applies random field theory (RFT) to identify significant clusters of vertices at the family-wise error rate (FWER) of p<0.05 within each subcortical structure, to account for the multiple comparisons inherent in surface maps [29]. Group differences were visualized as a color map displayed on the overall average surface [23].

We then extracted the significant vertices found in the TS-HC surface comparison in applicable structures and found the mean values for each subject of the deformation from the template at these vertices. Additionally, significant vertices from the NT-HC surface comparisons were studied with partial correlation analyses on each subject’s mean deformation value and their 12-month clinical score (TTS), while controlling for TTS at screening, age, and sex. Subsequently, we repeated this process using significant vertices we extracted from the NT-TS surface comparisons.

## Results

### Subject demographics and clinical features

In total, we analyzed 187 participants: 60 HC (44M / 16F), 88 NT (63M / 25F), and 39 TS (28M / 11F) (Table 1). Sex, mean age (all near 8 years old), scanner type used, and race did not differ significantly between groups.

**Table 1.** Participant characteristics at baseline or 12-months. (as specified under “Sample Characteristic”). Values indicate number or mean ± SD unless indicated otherwise.

| Sample Characteristic | HC | NT | TS | Statistics |
| --- | --- | --- | --- | --- |
| Total N (M / F) | 60 (44M / 16F) | 88 (63M / 25F) | 39 (28M / 11F) | $\chi^2 = 0.06, p = 0.97$ |
| Mean age (SD) at baseline (years) | 8.3 ± 2.0 | 7.8 ± 1.9 | 8.4 ± 1.9 | F=1.83, $p=0.16$ |
| Scanner Type | 15 Trio / 45 Prisma | 28 Trio / 60 Prisma | 8 Trio / 31 Prisma | $\chi^2 = 1.97, p = 0.37$ |
| Race | 44 White / 10 Black or African American / 5 More than one race / 1 Unknown or not reported | 73 White / 6 Black or African American / 6 More than one race / 1 Asian / 2 Unknown or not reported | 30 White / 8 More than one race / 1 Asian | $\chi^2 = 17.10, p = 0.07$ |
| Mean TTS at baseline | N/A | 16.96 ± 5.64; $n=80$ | 20.79 ± 7.22; $n=39$ | N/A |
| Mean TTS at 12 months | N/A | 14.16 ± 6.94; $n=80$ | N/A | N/A |

### Baseline TIV

The one-way ANOVA on TIV with age and sex as covariates revealed significant group differences [F(2,182)=3.316, p=0.004; see Fig. 1]. Post hoc Tukey HSD tests revealed that the NT group had smaller TIV than controls at baseline (NT=1484 ± 154 cm³ and HC= 1596 ± 141 cm³; p=0.03). Group differences in TIV between NT and healthy controls remained significant after controlling for both scanner type and race independently. Additionally, the TIV differed significantly across scanner types (p=0.04) and race (p=0.04). Since TIV differed significantly among groups, we included TIV as a factor when scaling the surfaces.

**Figure 1.**
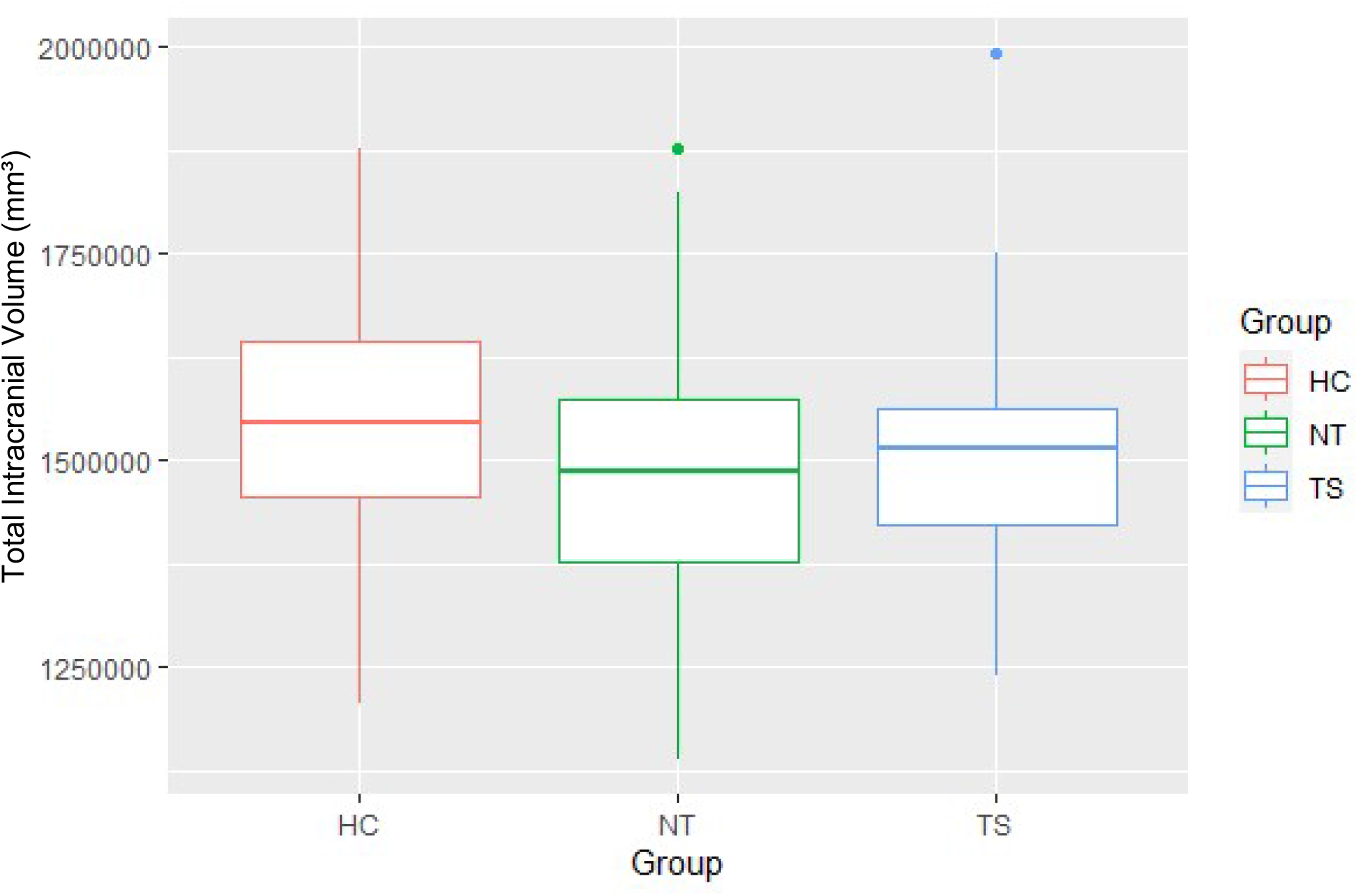
Group comparisons of total intracranial volume measured at baseline, accounting for age and sex. Healthy controls had the highest average, followed by TS and NT. ANCOVA analyses found an overall group effect (see above).

### Baseline group comparisons of subcortical structural volumes

Table 2 summarizes the estimated marginal means and standard errors of the subcortical volume for each group accounting for age and sex, as well as statistical comparisons for each group. Here, we describe the results for each structure in detail.

**Table 2.**
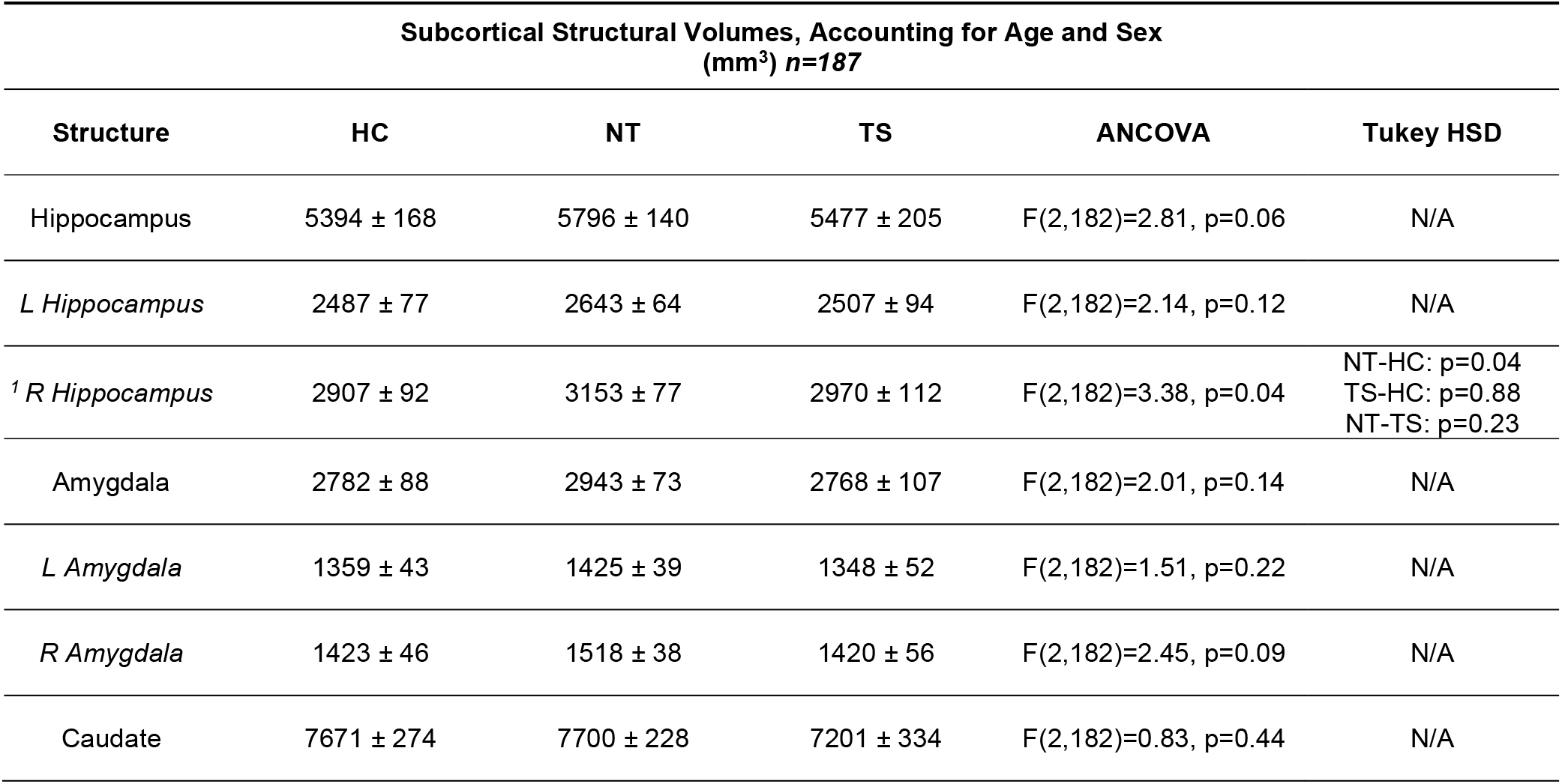
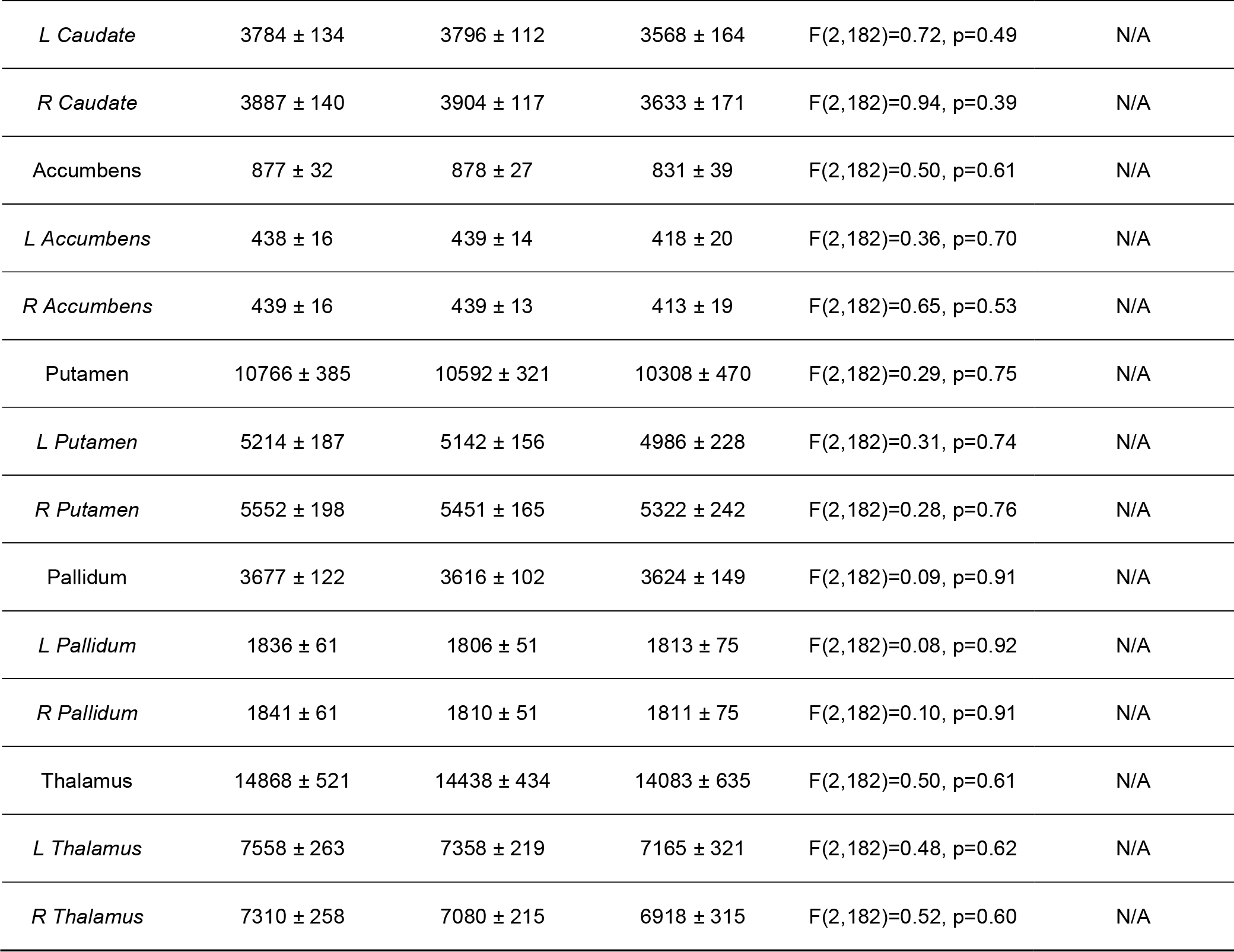
Subcortical Structural Volumes, Accounting for Age and Sex. Significant group differences were found in the hippocampus using a one-way ANCOVA while controlling for age and sex. Post hoc Tukey HSD tests further revealed a specific group difference: ^1^ NT had larger right hippocampal volumes than healthy controls (p = 0.04). Values indicate estimated marginal means ± SE unless indicated otherwise.

| Subcortical Structural Volumes, Accounting for Age and Sex<br>(mm³) *n*=187 |  |  |  |  |  |
| --- | --- | --- | --- | --- | --- |
| Structure | HC | NT | TS | ANCOVA | Tukey HSD |
| Hippocampus | 5394 ± 168 | 5796 ± 140 | 5477 ± 205 | F(2,182)=2.81, p=0.06 | N/A |
| *L Hippocampus* | 2487 ± 77 | 2643 ± 64 | 2507 ± 94 | F(2,182)=2.14, p=0.12 | N/A |
| <sup>1</sup> *R Hippocampus* | 2907 ± 92 | 3153 ± 77 | 2970 ± 112 | F(2,182)=3.38, p=0.04 | NT-HC: p=0.04<br>TS-HC: p=0.88<br>NT-TS: p=0.23 |
| Amygdala | 2782 ± 88 | 2943 ± 73 | 2768 ± 107 | F(2,182)=2.01, p=0.14 | N/A |
| *L Amygdala* | 1359 ± 43 | 1425 ± 39 | 1348 ± 52 | F(2,182)=1.51, p=0.22 | N/A |
| *R Amygdala* | 1423 ± 46 | 1518 ± 38 | 1420 ± 56 | F(2,182)=2.45, p=0.09 | N/A |
| Caudate | 7671 ± 274 | 7700 ± 228 | 7201 ± 334 | F(2,182)=0.83, p=0.44 | N/A |
| *L Caudate* | 3784 ± 134 | 3796 ± 112 | 3568 ± 164 | F(2,182)=0.72, p=0.49 | N/A |
| *R Caudate* | 3887 ± 140 | 3904 ± 117 | 3633 ± 171 | F(2,182)=0.94, p=0.39 | N/A |
| *Accumbens* | 877 ± 32 | 878 ± 27 | 831 ± 39 | F(2,182)=0.50, p=0.61 | N/A |
| *L Accumbens* | 438 ± 16 | 439 ± 14 | 418 ± 20 | F(2,182)=0.36, p=0.70 | N/A |
| *R Accumbens* | 439 ± 16 | 439 ± 13 | 413 ± 19 | F(2,182)=0.65, p=0.53 | N/A |
| *Putamen* | 10766 ± 385 | 10592 ± 321 | 10308 ± 470 | F(2,182)=0.29, p=0.75 | N/A |
| *L Putamen* | 5214 ± 187 | 5142 ± 156 | 4986 ± 228 | F(2,182)=0.31, p=0.74 | N/A |
| *R Putamen* | 5552 ± 198 | 5451 ± 165 | 5322 ± 242 | F(2,182)=0.28, p=0.76 | N/A |
| *Pallidum* | 3677 ± 122 | 3616 ± 102 | 3624 ± 149 | F(2,182)=0.09, p=0.91 | N/A |
| *L Pallidum* | 1836 ± 61 | 1806 ± 51 | 1813 ± 75 | F(2,182)=0.08, p=0.92 | N/A |
| *R Pallidum* | 1841 ± 61 | 1810 ± 51 | 1811 ± 75 | F(2,182)=0.10, p=0.91 | N/A |
| *Thalamus* | 14868 ± 521 | 14438 ± 434 | 14083 ± 635 | F(2,182)=0.50, p=0.61 | N/A |
| *L Thalamus* | 7558 ± 263 | 7358 ± 219 | 7165 ± 321 | F(2,182)=0.48, p=0.62 | N/A |
| *R Thalamus* | 7310 ± 258 | 7080 ± 215 | 6918 ± 315 | F(2,182)=0.52, p=0.60 | N/A |

#### Hippocampus

Groups differed in whole hippocampal volume at p=.06. The difference reached statistical significance for the right hippocampus (p=.04). NT participants had on average an 8.5% larger right hippocampus compared to the children without tics.

#### Amygdala, caudate, accumbens, putamen, pallidum, and thalamus

No group differences were found among HC, NT, and TS groups.

### Prediction of 12-month TTS from baseline structural volume

Longitudinal TTS analyses included 80 NT subjects. Over the course of 8.5 months (median) between the baseline and second visits, the average TTS decreased significantly from 16.96 ± 5.64 to 14.16 ± 6.94 (t = 3.82, df = 79, p-value = 0.0003).

Table 3 summarizes the partial correlation values between structural baseline volumes and TTS changes, while controlling for screen TTS, age, and sex. Here, we describe the results for each structure in detail.

**Table 3.** Partial Correlation between Baseline Volume and TTS. (controlled for screen TTS, age, & sex; n=80).

| **Partial Correlation between Baseline Volume and TTS Change**<br>(controlled for screen TTS, age, & sex; $n=80$) | | |
| --- | --- | --- |
| **Structure** | **TTS Change (r)** | **p-Value** |
| Hippocampus | 0.18 | 0.12 |
| Amygdala | 0.17 | 0.14 |
| Caudate | 0.21 | 0.06 |
| Accumbens | 0.15 | 0.20 |
| Putamen | 0.19 | 0.09 |
| Pallidum | 0.24 | 0.04 |
| Thalamus | 0.23 | 0.05 |
| TIV | -0.14 | 0.23 |

#### Hippocampus, amygdala, accumbens, putamen, and TIV

Scaled baseline structural volumes and TIV did not significantly predict TTS changes in the NT group. All correlations had r between -0.14 and 0.24.

#### Caudate

We found a positive correlation between baseline caudate volume and TTS change from baseline to 12-month measurements that was near significance (r=0.21, p=0.06).

#### Pallidum

We found a significant positive correlation between baseline pallidal volume and TTS change from baseline to 12-month measurements (r=0.24, p=0.04). In other words, a larger pallidum at baseline was significantly correlated with less improvement of tic symptoms.

#### Thalamus

We found a significant positive correlation between baseline thalamic volume and TTS change from baseline to 12-month measurements (r=0.23, p=0.05). In other words, a larger thalamus at baseline was significantly correlated with less improvement of tic symptoms.

### Baseline group comparisons of subcortical structural shape

When comparing the NT group to healthy controls, two structures were found to have significant regions of deformation as described below.

#### Putamen

(Figure 2 Panel A). Inferiorly, a small region of inward deformation was found in the right medial putamen.

**Figure 2.**
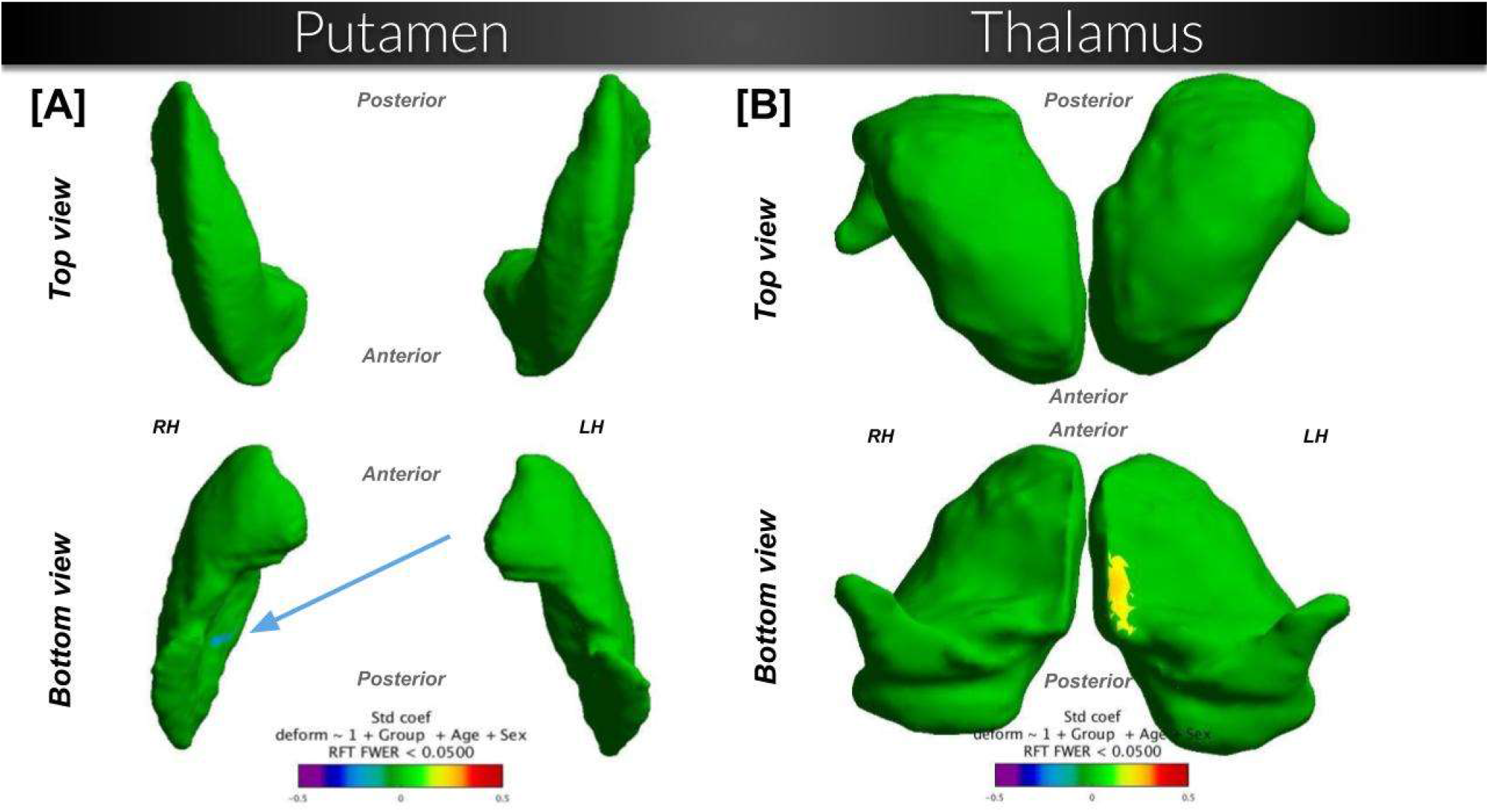
Shape comparison between NewTics vs. Healthy Controls. (n=148; 88 NT and 60 HC), while controlling for the effects of age and sex. Surfaces are scaled by total intracranial volume and voxel resolution. Cooler shades represent a greater inward deformation of the first group relative to the second, whereas warmer shades represent greater outward deformation. RFT = comparisons that passed the random field theory threshold.

#### Thalamus

(Figure 2 Panel B). We found a region of outward deformation in the medial aspect of the inferior left thalamus.

#### Hippocampus, amygdala, caudate, accumbens, and pallidum

No focal group shape differences were found among HC, NT, and TS groups.

Significant differences were also found comparing the NT group to TS children. In this comparison, six structures were found to have significant regions of deformation as described below.

#### Hippocampus

(Figure 3 Panel A). Superiorly, regions of outward deformation (NT > TS) are concentrated towards the lateral-posterior parts of both the left and right hippocampus.

**Figure 3.**
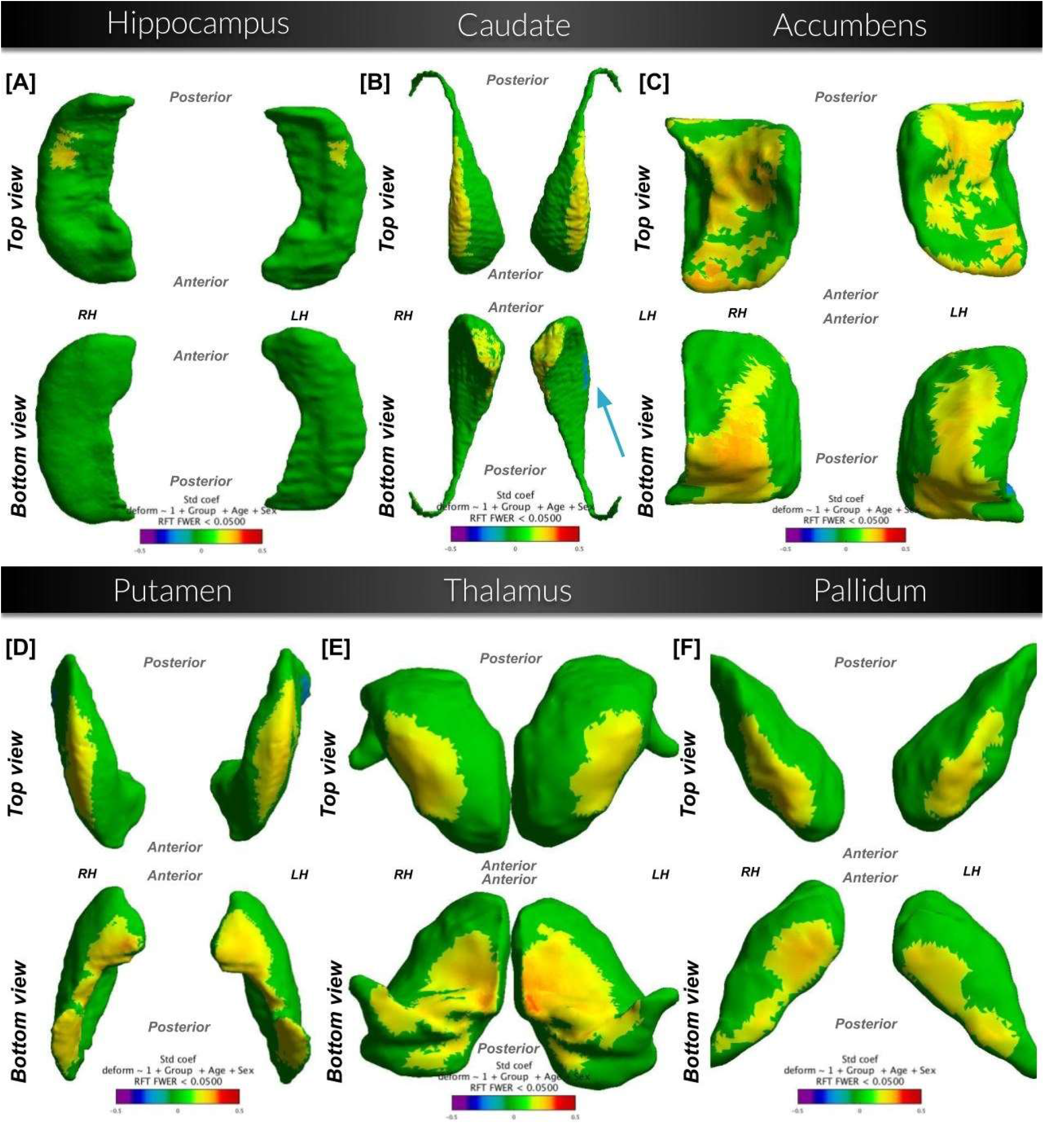
Shape comparison between NewTics vs. Tourette Syndrome. (n=127; 88 NT and 39 TS), while controlling for the effects of age and sex. Surfaces are scaled by total intracranial volume and voxel resolution. Cooler shades represent a greater inward deformation of the NT group relative to the TS group, whereas warmer shades represent greater outward deformation. RFT = comparisons that passed the random field theory threshold.

#### Amygdala

No group differences were found among HC, NT, and TS groups.

#### Caudate

(Figure 3 Panel B). Superiorly, large regions of outward deformation (NT > TS) can be seen along the lateral edge of both the left and right caudate. Inferiorly, regions of outward deformation are seen in the medial anterior parts of the left and right caudate. Additionally, a small region of inward deformation exists inferiorly along the lateral edge of the left caudate.

#### Accumbens

(Figure 3 Panel C). Several large regions of outward deformation (NT > TS) appear in both the left and right hemispheres superiorly and inferiorly.

#### Putamen

(Figure 3 Panel D). Inferiorly, a small region of inward deformation (NT > TS) is seen in the right medial putamen.

#### Thalamus

(Figure 3 Panel E). We found a region of outward deformation (NT > TS) in the medial left thalamus inferiorly.

#### Pallidum

(Figure 3 Panel F). We found regions of outward deformation (NT > TS) in both the left and right pallidum inferiorly and superiorly.

When comparing the TS group to healthy controls, four structures were found to have significant regions of outward deformation. In other words, areas of outward deformation described below illustrate localized volume gain in the specified regions in TS.

#### Hippocampus, amygdala, and pallidum

No group differences were found among HC, NT, and TS groups.

#### Caudate

(Figure 4 Panel A). Inferiorly, two small regions of outward deformation (TS > HC) are seen in both the left and right medial caudate.

**Figure 4.**
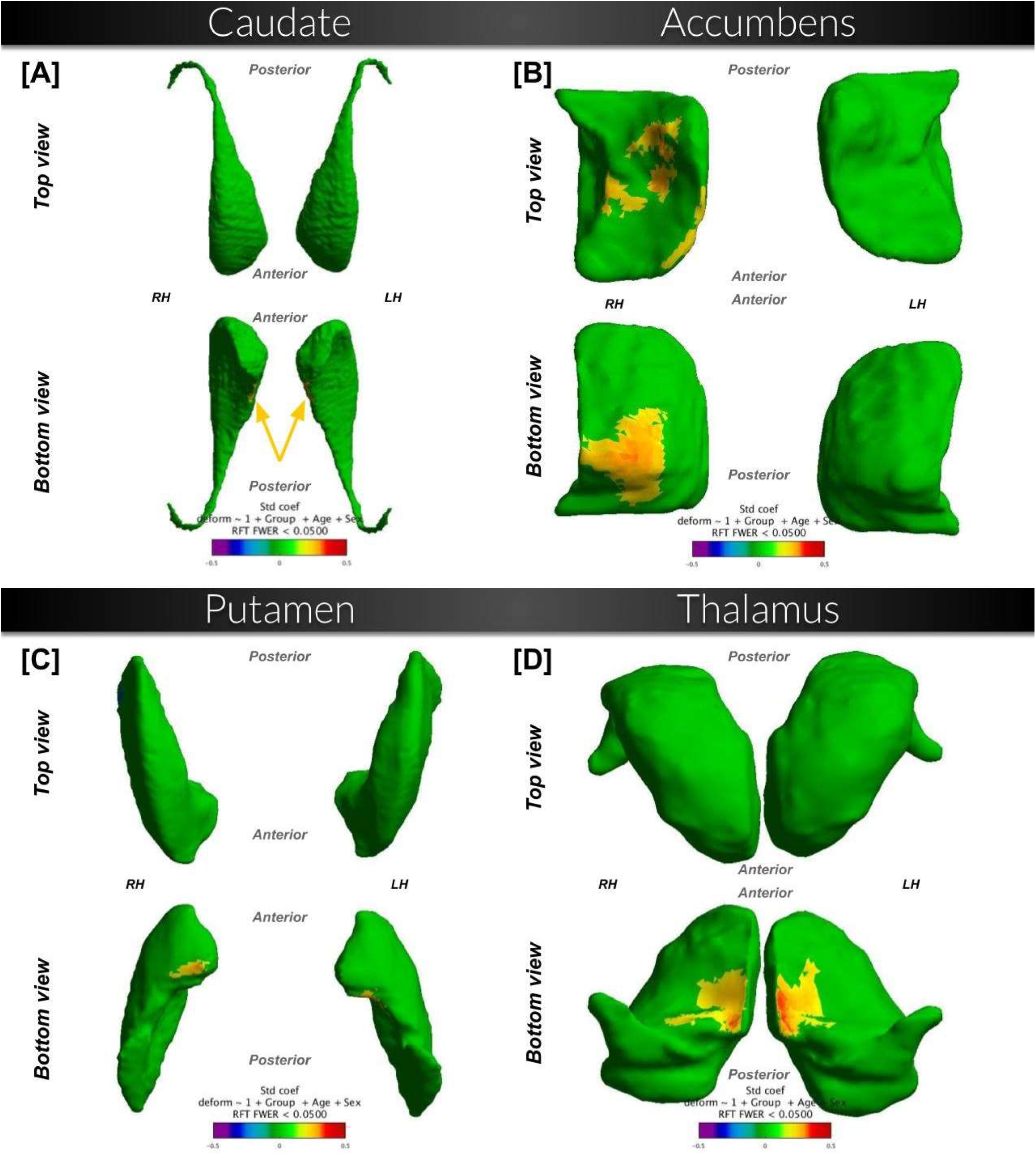
Shape comparison between Tourette Syndrome vs. Healthy Controls. (n=99; 29 TS and 60 HC), while controlling for the effects of age and sex. Surfaces are scaled by total intracranial volume and voxel resolution. Cooler shades represent a greater inward deformation of the first group relative to the second, whereas warmer shades represent greater outward deformation. RFT = comparisons that passed the random field theory threshold.

#### Accumbens

(Figure 4 Panel B). Superiorly and inferiorly, regions of outward deformation can be seen in the right accumbens.

#### Putamen

(Figure 4 Panel C). Inferiorly, small regions of outward deformation are seen in the left and right medial anterior putamen.

#### Thalamus

(Figure 4 Panel D). We found regions of outward deformation towards the medial ends of the left and right thalamus.

Table 4 summarizes the mean values for each group of the deformation from the template within the significant vertices found in the TS-HC surface comparison, as well as statistical comparisons for each group. Here, we describe the results for each structure in detail.

**Table 4.** Deformation from Template Group Values. Significant group differences were found in the caudate using a one-way ANCOVA while controlling for age and sex. Post hoc Tukey HSD tests further revealed a specific group difference: ^1^ In the caudate, TS demonstrated overall outward deformation from the template in significant vertices compared to the overall inward deformation seen in healthy controls (p = 0.03). ^2^ In the caudate, TS also demonstrated overall outward deformation from the template in significant vertices compared to the overall inward deformation seen in NT (p = 0.02). Values indicate mean deformation value unless indicated otherwise

| Deformation from Template Group Values<br>*n=187* |  |  |  |  |  |
| --- | --- | --- | --- | --- | --- |
| Structure | HC | NT | TS | ANCOVA | TukeyHSD |
| Caudate | -0.0489 | -0.0510 | 0.1903 | F(2,182)=4.39, p=0.01 | <sup>1</sup> TS-HC: p=0.03<br><sup>2</sup> TS-NT: p=0.02 |
| Accumbens | -0.0611 | 0.0066 | 0.0792 | F(2,182) =1.91, p=0.15 | N/A |
| Putamen | -0.0222 | -0.0158 | 0.0698 | F(2,182)=0.82, p=0.44 | N/A |
| Thalamus | -0.0350 | -0.0039 | 0.0627 | F(2,182)=0.45, p=0.64 | N/A |
| All significant vertices | -0.0465 | -0.0116 | 0.0977 | F(2,182)=1.85, p=0.16 | N/A |

#### Caudate

We found overall average outward deformation compared to the template in the TS group, compared to inward deformation compared to the template in both the HC and NT groups.

#### Accumbens, putamen, thalamus, and all significant vertices as a whole

there were no statistically significant differences between groups.

#### Prediction of 12-month TTS from baseline structural shape

When comparing the NT group with healthy controls, we found that 18 out of the 13638 vertices on the putamen surface had a significant deformation value. On the thalamus, 45 out of 9580 vertices were significant.

Next, we tested whether these focal deformations at baseline predicted clinical change at follow-up. Table 5 summarizes the correlation and p-values between each NT subject’s mean deformation value in the significant vertices of the NT-HC comparison and their 12-month TTS, while controlling for screen TTS, age, and sex. We did not find a significant correlation between mean deformation values and 12-month TTS.

**Table 5.**
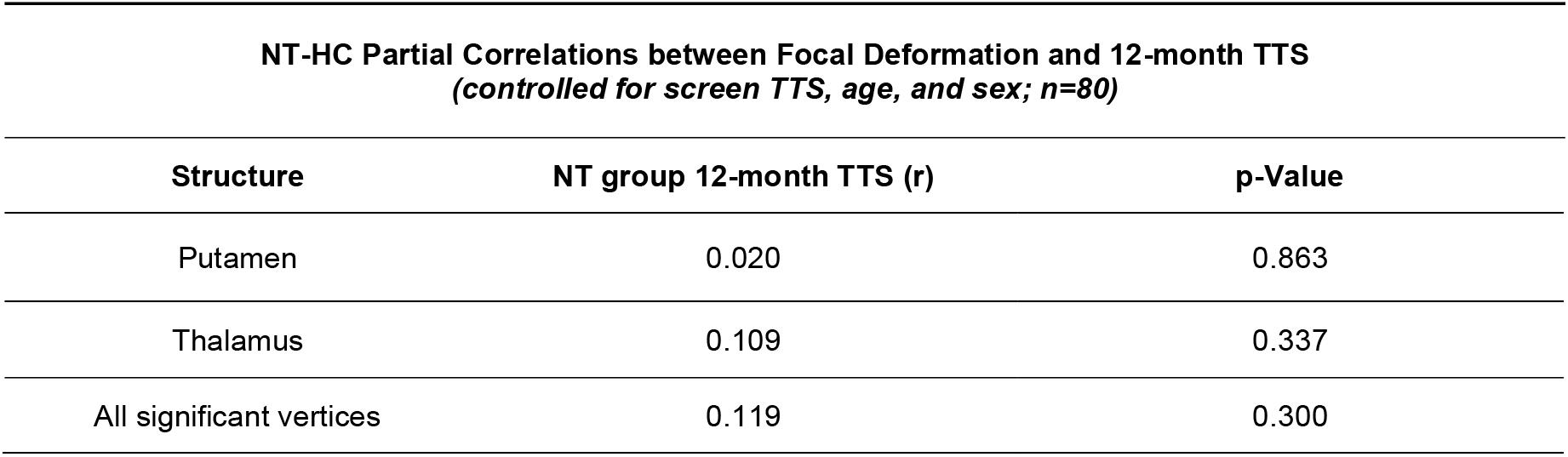
NT-HC Partial Correlations between Deformation and 12-month TTS. (controlled for screen TTS, age, and sex; n=80).

| **NT-HC Partial Correlations between Focal Deformation and 12-month TTS**<br>**(controlled for screen TTS, age, and sex; n=80)** |  |  |
| --- | --- | --- |
| **Structure** | **NT group 12-month TTS (r)** | **p-Value** |
| Putamen | 0.020 | 0.863 |
| Thalamus | 0.109 | 0.337 |
| All significant vertices | 0.119 | 0.300 |

When comparing the NT group with the TS group, numbers of significant vertices on structure surfaces were as follows:183/13222 significant vertices on the hippocampus surface; 3734/24744 significant vertices on the caudate surface; 2279/5804 significant vertices on the nucleus accumbens surface; 1030/5398 significant vertices on the pallidum surface; 3345/13638 significant vertices on the putamen surface; 1956/9580 significant vertices on the thalamus surface.

Table 6 summarizes the correlation and p-values between each NT subject’s mean deformation value in the significant vertices of the NT-TS comparison and their 12-month TTS, while controlling for screen TTS, age, and sex. We did not find a significant correlation between mean deformation values and 12-month TTS.

**Table 6.** NT-TS Partial Correlations between Deformation and 12-month TTS. (controlled for screen TTS, age, and sex; n=80).

|  |  |  |
| --- | --- | --- |
| **Structure** | **NT group 12-month TTS (r)** | **p-Value** |
| Hippocampus | -0.101 | 0.374 |
| Caudate | -0.125 | 0.274 |
| Accumbens | -0.092 | 0.421 |
| Pallidum | -0.135 | 0.235 |
| Putamen | -0.145 | 0.204 |
| Thalamus | -0.144 | 0.206 |
| All significant vertices | -0.133 | 0.242 |

## Discussion

In this study, we focus on the first year of tic development (NT group) and identified several baseline subcortical volume and shape characteristics related to baseline tic symptoms and some that predict clinical tic outcome 3 to 12 (mean 7.9) months later.

Baseline volume analyses showed that the right hippocampus was on average 8.5% larger in NT children compared to healthy controls. Left hippocampus was also larger (6.3%), but non-significantly (p=.12). A larger pallidum or thalamus at baseline predicted a weaker improvement of tic symptoms between at follow-up.

Surface analyses demonstrated distinct patterns of subcortical surface deformation in several structures across all group comparisons. When comparing the NT group to healthy controls, we found that the putamen exhibited inward deformation (i.e., localized volume loss), while the thalamus showed outward deformation (localized volume gain). In the TS group, each of the studied structures except amygdala showed significant outward deformation compared to healthy controls. Similarly, this pattern was present when comparing the NT group to the TS group, with the NT group showing consistent patterns of greater outward deformation in the caudate, accumbens, putamen, and thalamus. However, none of the NT-vs.-HC or NT-vs.-TS deformation values significantly predicted 12-month TTS.

Finally, we compared each participant group to the template. In the significant vertices from the caudate TS-HC surface comparison, the TS group showed an overall trend of outward deformation from the template compared to both the HC and NT groups. Each NT subject’s mean deformation value at the NT-HC or NT-TS significant vertices for any region did not significantly predict their 12-month TTS.

### Functional implications

#### Hippocampus

Children in the NT group had larger right hippocampal volume compared to healthy controls. The NT group also exhibited localized volume increases compared to the TS group. The hippocampus plays a role in memory consolidation in both the cognitive and motor domains [30]. Evidence is strong for a relationship with visuospatial memory, with a larger hippocampus associated with higher performance and traumatic injury to the right hippocampus correlated to lower memory performance [31, 32]. Our conjecture is that enlargement of the hippocampus may lead to abnormally strong preservation of motor memory. In children with TS, those with more persistent visuomotor memory show more severe tics and took longer to unlearn a previously learned motor pattern [33]. As the NT group had a larger right hippocampus at baseline compared to healthy controls, we hypothesize that this region may be implicated in tic development and/or persistence. Previous work has found a larger hippocampus in children with *chronic* tics (TS) [34], but the current results, confirming our results with a smaller sample and a different method [16], show that the larger hippocampus is present very early in the course of tic disorder and cannot result from adaptation to years of ticcing. This conclusion is consistent with a study of almost 5,000 people with TS, which associated genetic risk variants for TS with gene variants linked to a larger hippocampus [35].

#### Basal Ganglia (accumbens, pallidum, putamen, and caudate)

A larger baseline pallidum predicted a weaker improvement of tic symptoms. When analyzing shape differences, we found significant patterns of localized volume increases in NT children compared to TS children in all four structures. However, the putamen in the NT group exhibited localized volume loss when compared to healthy controls, but this deformation was concentrated towards the posterior end. The TS group exhibited localized volume increases in the caudate, accumbens, and putamen when compared to healthy controls. In the significant vertices from the TS-HC surface comparison of the caudate, we found an overall trend of greater outward deformation (compared to the template) in the TS group compared to both the HC and NT groups.

We had hypothesized that smaller caudate volume would predict worse outcome (higher TTS scores at 12 months) based on previous findings [10, 14]. For example, a smaller caudate in 43 children with TS was found to predict more severe tic symptoms an average of 7.5 years later [36]. However, in the NT group, we found the opposite at p=0.06 (Table 3). Similarly, we had hypothesized that caudate volume would be significantly lower in NT when compared to healthy controls, though our findings suggest that caudate volume in NT was similar or slightly higher compared to controls (Table 2). Our contrasting results could be a result of exploring a newly studied population (our NT group) rather than only children with established TS in previous studies. Alternatively, the smaller caudate in the older work in TS [10] may have been an artifact attributable to greater head movement, and more parsimoniously attributed to more head movement in those with tics at the time of the scan. The present work has the advantage of prospective movement correction, minimizing such potential confounding.

The basal ganglia are subcortical nuclei that, along with their associated connections, have been a large focus of TS/CTD research. They are involved in motor control in other movement disorders such as Parkinson’s and Huntington’s disease. Cortico-striatal-thalamo-cortical (CSTC) circuits are involved in inhibitory control and habit formation, both of which are affected in TS [37, 38]. This understanding may relate to the shape and volume differences we found in basal ganglia structures and the pallidum’s association with observed changes in tic symptoms.

#### Thalamus

Thalamus volume did not differ significantly between groups. When analyzing surfaces, however, we found that the thalamus displayed localized volume increases in NT children when compared to both TS children and healthy controls. Previous volume and surface morphology studies have found greater grey matter volume or outward deformation in the thalamus of TS children compared to healthy controls, which is again seen here in our surface comparison between TS and healthy children [13, 39]. Further, we found a significant correlation between a larger thalamus at baseline and the subsequent lesser improvement in tic symptoms. The position of the thalamus in CSTC circuits, mediating output from motor-related basal ganglia and motor areas of the cerebral cortex, supports its involvement in motor control [40]. Outward surface deformation or increased volume in the medial thalamus may reflect a pathological process that leads to greater persistence of tics. As one possible mechanism, possibly pruning of local synaptic connections in healthy development reduces the risk of tic persistence, and deficiencies in that pruning help reinforce tics.

Our results are also consistent with the results of a meta-analysis on task-based fMRI studies in patients with TS [41]. In the thalamus, the meta-analysis showed a clear overlap of all the conditions involving various aspects of voluntary motor execution, response inhibition, and tic generation. The pallidum and thalamus both showed more consistent activation in free-to-tic conditions. The meta-analysis also identified a positive correlation between tic severity and BOLD activity in the thalamus and putamen [41]. Further, another study using whole brain diffusion tensor imaging (DTI) found alteration in the thalamus and putamen, congruent with other DTI studies in adult TS patients [42]. Similarly, in the putamen and thalamus, we found that the NT group exhibited localized volume gain compared to the TS group, and the TS group showed localized volume gain compared to the healthy controls. It seems that these regions are implicated in both tic severity and persistence. In the thalamus, most of patterns of outward deformation seen in the NT and TS comparisons with healthy controls (Figure 2 and Figure 4) were specifically found in the mediodorsal thalamus [43, 44]. This region in the thalamus has become one of the most often targeted regions for deep brain stimulation in patients with treatment-resistant tic disorders [45].

Further, when considering a volume increase-decrease model, we can perhaps consider the NT and TS groups to be different timepoints on the same timeline. In this case, the strongest group differences indicated by larger subcortical volumes and outward deformation are seen in NT children when compared to TS and HC groups, and these results either normalize or reverse as the tics persist or disappear over time. A similar pattern was found when studying children and adults with autism spectrum disorder (ASD) compared with healthy participants. Compared with control participants, patients with ASD showed the strongest group differences (increased thickness in the frontal areas and decreased thickness in the temporal lobe) during childhood and adolescence, with normalized or even reversed thickness results in adulthood [46]. These results suggested a complex developmental course for frontal, temporal, and subcortical structures in ASD. When comparing children and adults with TS, hippocampus and amygdala volumes declined significantly with age in the TS group but not in controls [34]. In the case of TS, the subcortical enlargement seen in the NT group could possibly delay onset of typical developmental processes and then subsequent normalization occurs, such that chronic cases (TS/CTD) exhibit less abnormality than those with newly developed tics when compared to healthy controls.

One limitation of this project is that we currently lack 12-month clinical data for a small fraction of the NT participants for whom we have baseline scan data, so we were somewhat limited in which participants we could include in predictive analyses. Additionally, our healthy controls were collected across different studies/sources. While we performed harmonization steps (such as adjusting for voxel size differences), they were still not all from the same cohort, and thus had slightly varying scanning parameters. We had also planned in advance to correct for differences in TIV, and the results showing significantly smaller TIV in the NT group confirmed the need for that plan. However, if all regional volumes in the NT group differ in the same direction from the other groups, one wonders whether the apparent regional changes simply reflect the whole-brain difference. However, even without correction for TIV, the hippocampal volume is still numerically larger in the NT group, taking age and sex into account.

Nevertheless, the New Tics study does provide the first imaging results from Provisional Tic Disorder. These new findings have potential clinical relevance, such as strengthening the possibility to identify ideal targets for treatment optimization. Continuing to investigate neuroanatomical characteristics in Provisional Tic Disorder may also further provide insight into prognostic biomarkers. In many children with Provisional Tic Disorder, tics improve within the first year, often to the point of clinical insignificance, but previously there was a paucity of information to predict which children would instead go on to have more severe tics over time. Understanding the mechanisms related to these outcomes may provide clinical insight into the pathophysiological traits in this complex disease and thus potentially guide interventions such as conventional or adaptative deep brain stimulation in drug-resistant cases, leading to an individualized treatment. A recent meta-analysis reports that deep brain stimulation in TS leads to a 40% improvement on tic severity scales on average [47]. However, given the invasiveness of deep brain stimulation, predictors of prognosis are urgently needed to direct deep brain stimulation only to those children least likely to improve spontaneously. Finally, with a deeper understanding of neural networks related to the recent development of tics and progression of symptoms, we may be able to provide more ideal targets that could be manipulated to prevent the worsening of tic symptoms.

## Data Availability

All data from the New Tics study will be available from the NIMH Data Archive (https://nda.nih.gov, study ID C2692).

## Author Contributions

Conceptualization, K.J.B., B.L.S. and L.W.; methodology, L.W.; formal analysis, T.C., L.W., J.D.; investigation, K.J.B., D.J.G., B.L.S., S.K., and T.H.; data curation, K.J.B. and S.K.; writing—original draft preparation, T.C.; writing—review and editing, all authors; visualization, T.C.; project administration, K.J.B.; funding acquisition, K.J.B., D.J.G., T.H., B.L.S. All authors have read and agreed to the published version of the manuscript.

## Acknowledgements

We thank Rebecca Coalson, Jessica Church, Jonathan Koller, and Judith Lieu for comparison MRI data from children with Tourette syndrome or with no tics.

## Funding

Research reported in this publication was supported by the National Institutes of Health: National Institute of Mental Health under award number K24MH087913 to K.J.B.; K01MH104592 to D.J.G.; R01MH104030 to K.J.B. and B.L.S.; R01MH118217 to D.J.G.; National institute of Neurological Disorders and Stroke R21NS091635 to B.L.S. and K.J.B.; NIDDK award R01DK064832 to T.H.; NICHD award R01HD070855 to T.H.; the Washington University Institute of Clinical and Translational Sciences grants UL1RR024992 and UL1TR000448; the Eunice Kennedy Shriver National Institute of Child Health and Human Development of the National Institutes of Health under Award Number U54HD087011 to the Intellectual and Developmental Disabilities Research Center at Washington University, and K23DC006638 to J.E.C. Lieu; the Mallinckrodt Institute of Radiology MIR-IDDRC Pilot Study Fund; the McDonnell Center for Systems Neuroscience; the National Institute of Biomedical Imaging and Bioengineering grant R01 EB020062 to L.W.; the National Science Foundation grants 1734853 and 1636893 to L.W. The studies presented in this work were carried out in part in the East Building MR Facility of the Washington University Medical Center. The content is solely the responsibility of the authors and does not necessarily represent the official views of the National Institutes of Health or of the MIR.

## Institutional Review Board Statement

Protocols 201109157 and 201707059 were approved by the Washington University Human Research Protection Office (IRB).

## Informed Consent Statement

Each child assented, and a parent (guardian) gave informed consent. Data shared from other projects were shared after appropriate human subjects review and consent.

## Conflicts of Interest

The authors declare no conflicts of interest.

## Notes

### Competing Interest Statement

The authors have declared no competing interest.

### Author Declarations

The work described herein was approved by the Washington University Human Research Protection Office (IRB), protocol numbers 201109157 and 201707059. Each child assented and a parent (guardian) gave informed consent. Data shared from other projects were shared after appropriate human subjects review and consent.

### Summary of Updates

Added author Tamara Hershey for her contribution with data collection; included baseline TTS for the TS group; reworded and elaborated further in some sections; added more literature reviews

